# RNAseq of amniotic fluid cell-free RNA: insights into the pathophysiology of congenital cytomegalovirus infection

**DOI:** 10.1101/2022.01.18.21268443

**Authors:** Lisa Hui, Luc De Catte, Sally Beard, Jovana Maksimovic, Neeta L Vora, Alicia Oshlack, Susan P Walker, Natalie J Hannan

**Affiliations:** Department of Obstetrics and Gynaecology, University of Melbourne, Victoria, Australia; Department of Obstetrics and Gynaecology, Mercy Hospital for Women, Heidelberg, Victoria Australia; Reproductive Epidemiology group, Murdoch Children’s Research Institute Parkville, Victoria Australia; Dept of Obstetrics and Gynaecology, Northern Health, Epping, Victoria Australia; Department of Gynaecology and Obstetrics, Universitair Ziekenhuis, Leuven, Belgium; Computational Biology group, Peter MacCallum Cancer Centre, Parkville, Victoria Australia; Department of Paediatrics, University of Melbourne, Victoria, Australia; Respiratory group, Murdoch Children’s Research Institute, Parkville, Victoria, Australia; Department of Obstetrics and Gynaecology, University of North Carolina at Chapel Hill, Chapel Hill, USA; Department of Biosciences, University of Melbourne, Victoria, Australia

**Keywords:** congenital CMV, transcriptomics, amniotic fluid, fetal neurodevelopment, cell-free RNA, RNA sequencing, RNAseq, cytomegalovirus, perinatal infections, biomarkers

## Abstract

**Background:** Congenital cytomegalovirus (cCMV) is the most common perinatal infection and a significant cause of sensorineural hearing loss, cerebral palsy and neurodevelopmental disability. There is a paucity of human gene expression studies examining the pathophysiology of CMV infection.

**Objectives:** The aim of this study was to perform a whole transcriptomic assessment of amniotic fluid from pregnancies with live fetuses to identify differentially-expressed genes and enriched Gene Ontology categories associated with cCMV infection.

**Study design:** Amniotic fluid supernatant was prospectively collected from pregnant women undergoing amniocentesis for suspected cCMV due to first trimester maternal primary infection or ultrasound features suggestive of fetal infection. Women who had received therapy to prevent fetal infection were excluded. cCMV was diagnosed via viral PCR of amniotic fluid; CMV-infected fetuses were paired with noninfected controls, matched for gestational age and fetal sex. Paired-end RNA sequencing was performed on amniotic fluid cell-free RNA with the Novaseq6000 at a depth of 30 million reads/sample. Following quality control and filtering, reads were mapped to the human genome and counts summarised across genes. Differentially expressed genes were identified using two approaches: voomWithQualityWeights in conjunction with *limma* and *RUVSeq* with *edgeR*. Genes with a false discovery rate (FDR) < 0.05 were considered statistically significant. Differential exon usage was analysed using *DEXSeq*. Functional analysis was performed using Gene Set Enrichment Analysis and Ingenuity Pathway Analysis. Manual curation of differentially regulated genes was also performed.

**Results:** Amniotic fluid samples were collected from 50 women; 16 (32%) had cCMV confirmed by PCR. After excluding 3 samples without matched controls, 13 CMV-infected samples collected at 18-23 weeks and 13 CMV-negative gestation matched controls were submitted for RNA sequencing and analysis (total n=26). Ten of the 13 pregnancies with CMV-infected fetuses had amniocentesis due to serological evidence of maternal primary infection with normal fetal ultrasound, and three had amniocentesis due to ultrasound abnormality suggestive of CMV infection. Four CMV-infected pregnancies ended in termination (n=3) or fetal death (n=1), and 9 resulted in live births. Pregnancy outcomes were available on 11 of the 13 CMV-negative controls; all resulted in live births of clinically-well infants.

Differential gene expression analysis revealed 309 up-regulated and 32 down-regulated genes in the CMV-infected group compared with the CMV-negative group. Gene set enrichment analysis showed significant enrichment of multiple Gene Ontology categories involving the innate immune response to viral infection and interferon signalling. Of the 32 significantly down-regulated genes, 8 were known to be involved in neurodevelopment and preferentially expressed by the brain. Six specific cellular restriction factors involved in host defence to CMV infection were upregulated in the CMV-infected group. Ingenuity Pathway Analysis predicted activation of pathways involved in ‘progressive neurological disease’, and ‘inflammatory neurological disease’.

**Conclusions:** In this next generation sequencing study, we reveal new insights into the pathophysiology of cCMV infection. These data on the upregulation of the intraamniotic innate immune response to CMV infection and the dysregulation of neurodevelopmental genes may inform future approaches to developing prognostic markers and assessing fetal responses to in utero therapy.

## Introduction

Congenital CMV is the leading infectious cause of congenital malformations, stillbirth, sensorineural hearing loss and neurodevelopmental impairment in developed countries.^1^ It is also a major preventable cause of cerebral palsy, implicated in 10% of affected children.^2^ The fetal brain is especially vulnerable to injury from CMV due to the viral tropism for neuroprogenitor cells, mediated by both the direct cytotoxic effects of viral replication and bystander damage due to inflammation and microglial cell activation.^3^ While a prenatal diagnosis of congenital CMV can be made by viral DNA detection in amniotic fluid, prediction of long term neurodevelopment outcomes for infected fetuses is more challenging.

There are well established fetal brain abnormalities associated with cCMV, and destructive lesions are strong predictors for adverse outcome. However, normal fetal imaging in an infected fetus cannot exclude important outcomes such as hearing loss, or neurodevelopmental delay. Indeed, about half of children with postnatal sequelae have normal fetal imaging. ^4^ There have been numerous non-imaging parameters investigated for predicting outcome using maternal blood, amniotic fluid (AF) or fetal blood, but none have yet achieved reliable clinical utility.^5,6^

There is a paucity of human gene expression studies on the pathophysiology of CMV infection, mainly due to the lack of access to fresh tissue samples. A novel and feasible approach to studying the impact of CMV infection on human development and identifying candidate biomarkers of abnormal neurodevelopment is to analyse amniotic fluid RNA. ^7,8^ Cell-free RNA (cfRNA) in AF supernatant is derived from a range of fetal organs, including fetal brain. A number of investigators have now demonstrated characteristic gene expression profiles using microarray or RNA sequencing assessing pregnancies with twin-twin transfusion syndrome, spina bifida, as well as normal preterm and term pregnancies. ^9– 12^ However, AF cfRNA has not been previously employed to study the impact of cCMV on fetal development.

The aim of this study was to perform a comparative transcriptomic study of AF from second trimester human fetuses to identify differentially-expressed genes (DEGs) and enriched Gene Ontology (GO) categories associated with in utero infection. We hypothesized that inflammation and neurodevelopmental processes would be significantly dysregulated in CMV-infected fetuses compared with non-infected fetuses.

## Materials and Methods

### Study design

This was a prospective clinical study recruiting women undergoing clinically-indicated amniocentesis for suspected fetal CMV infection. CMV-infected cases were defined as those with a positive amniotic fluid PCR result for CMV DNA, as performed by the local clinical diagnostic laboratory, and controls were pregnancies with amniotic fluid that tested PCR-negative for CMV. Sample size calculation details are available in Supplemental file 1.

### Subjects and samples

Central study coordination, laboratory work, and analysis were performed within the Department of Obstetrics and Gynaecology, University of Melbourne, Australia. Prospective recruitment of pregnant women with suspected CMV infection was conducted during 2017-2019 in two sites: the fetal medicine units at University Hospitals Leuven, Belgium, and the Mercy Hospital for Women, Melbourne, Australia. Fresh amniotic fluid samples were obtained from women undergoing clinically-indicated second trimester amniocentesis for suspected fetal infection with CMV after informed consent for research participation. Women who had serological evidence of maternal primary infection in early pregnancy, and/or for fetal ultrasound abnormalities suggestive of CMV infection, were eligible for inclusion. Women who had received prenatal therapy for prevention of fetal infection with CMV hyperimmune globulin or antiviral medications such as valaciclovir were excluded. AF was sent for clinical diagnostic testing for CMV, and an aliquot of AF supernatant was retained for research after being centrifuged at 300 x *g* for 10 minutes within 6 hours of collection. Samples from Leuven were transported in a temperature-controlled manner to the University of Melbourne, Department of Obstetrics and Gynaecology, on dry ice. The amniotic fluid supernatant was stored at -80°C.

Due to slower-than-expected recruitment, a subset of amniotic fluid supernatant samples was obtained from the biobank of the University of North Carolina at Chapel Hill in order to reach the sample size of 12 matched pairs. CMV-infected amniotic fluid samples were identified via a search query of the biobank database. CMV-negative samples in structurally-normal, euploid fetuses matched for gestational age and fetal sex were also provided from the biobank as paired controls. Clinical classification of fetal sex by ultrasound was confirmed by *Xist* expression on RNA sequencing.

### Laboratory methods

Total RNA from AF supernatant was extracted as previously described, using the Qiagen Circulating Nucleic Acid kit with the manufacturer’s recommended in-solution DNAase step, followed by concentration and clean up with the Qiagen MinElute Clean Up kit.^8^ RNA was eluted in 14ul of nuclease-free water. Quality control of the RNA was performed with the Agilent TapeStation system (TapeStation Analysis Software A.02.02 (SR1) © Agilent Technologies, Inc. 2017). Library preparation was performed with NuGEN Ovation SoLo RNA-seq kit (Tecan Genomics, Inc., Redwood City, USA) according to the manufacturer’s instructions.

### Differential gene expression analysis

Paired-end RNA sequencing was performed on the Novaseq 6000 at 30 million reads per sample. Detailed methods section with full descriptions of the differential gene expression, functional analysis with Gene Set Enrichment Analysis (GSEA) and Ingenuity Pathway Analysis (IPA), and differential exon usage analysis are provided in Supplemental file 1.

### Institutional approvals

Prospective approval for this study was obtained from human research ethics committee of Mercy Health (R16-24) on 27 June 2016. All participants provided written informed consent.

## Results

### Clinical characteristics of the study group

Amniotic fluid supernatant was obtained from 50 pregnant women between 2017-2019, of whom 32% (n=16) were confirmed to have fetal infection on amniotic fluid CMV PCR. Three CMV infected samples were excluded from analysis due the absence of matched controls. We therefore performed RNA sequencing on a final group of 13 CMV-infected and 13 CMV-negative samples at 18-22 weeks gestation, matched for fetal sex and gestational age (+/-2 weeks). All except one of the 13 matched pairs were also matched for recruitment site.

Clinical details of the study cohort are shown in table 1. Twenty-four women had amniocentesis performed for suspected fetal CMV infection based on positive maternal serology in early pregnancy (n=21) and fetal ultrasound abnormality (n=3). An additional two samples performed for other indications in fetuses without structural or chromosomal anomalies were provided by the UNC biobank as matched negative controls. None of the participants received fetal therapy with anti-viral medication or CMV hyperimmune globulin prior to amniocentesis. Four of the CMV infected fetuses ended in termination (n=3) or fetal death (n=1) and three live births had subsequent brain abnormalities on MRI and/or ultrasound.

**Table 1.**
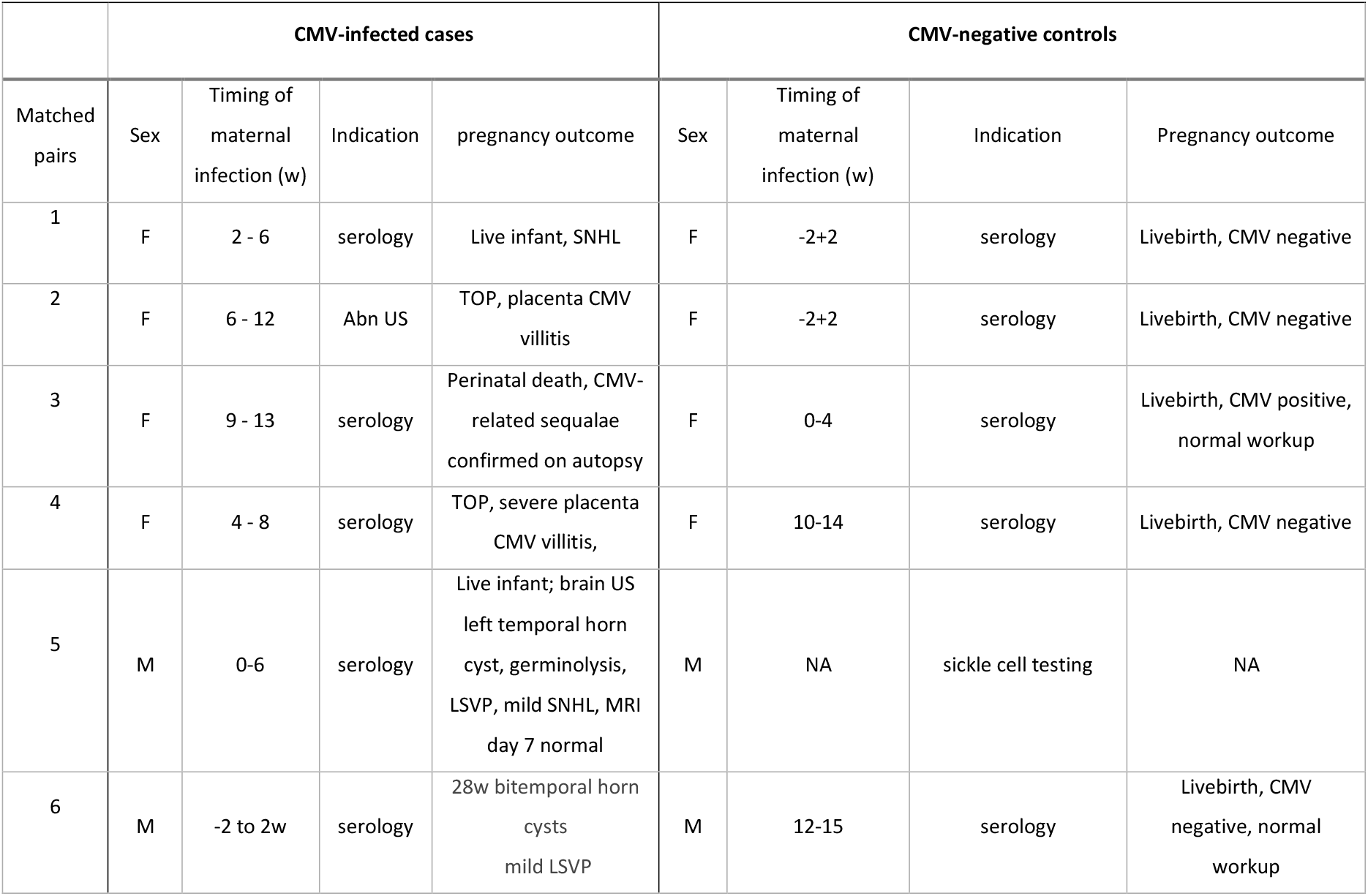
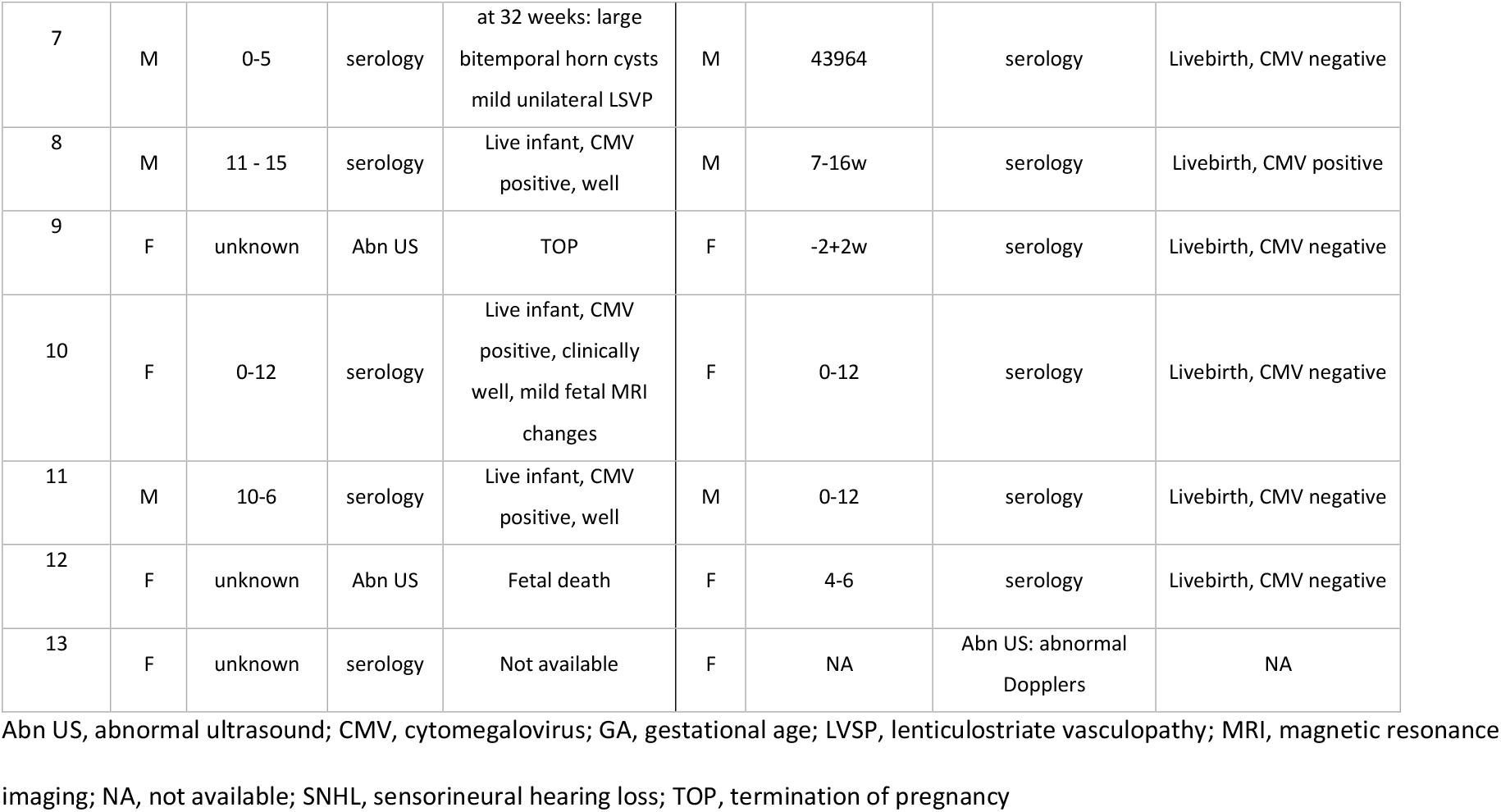
Clinical characteristics of CMV-infected cases and matched controls

|  | CMV-infected cases |  |  |  | CMV-negative controls |  |  |  |
| --- | --- | --- | --- | --- | --- | --- | --- | --- |
| Matched pairs | Sex | Timing of maternal infection (w) | Indication | pregnancy outcome | Sex | Timing of maternal infection (w) | Indication | Pregnancy outcome |
| 1 | F | 2 - 6 | serology | Live infant, SNHL | F | -2+2 | serology | Livebirth, CMV negative |
| 2 | F | 6 - 12 | Abn US | TOP, placenta CMV villitis | F | -2+2 | serology | Livebirth, CMV negative |
| 3 | F | 9 - 13 | serology | Perinatal death, CMV-related sequelae confirmed on autopsy | F | 0-4 | serology | Livebirth, CMV positive, normal workup |
| 4 | F | 4 - 8 | serology | TOP, severe placenta CMV villitis, | F | 10-14 | serology | Livebirth, CMV negative |
| 5 | M | 0-6 | serology | Live infant; brain US left temporal horn cyst, germinolysis, LSVP, mild SNHL, MRI day 7 normal | M | NA | sickle cell testing | NA |
| 6 | M | -2 to 2w | serology | 28w bitemporal horn cysts mild LSVP | M | 12-15 | serology | Livebirth, CMV negative, normal workup |
| 7 | M | 0-5 | serology | at 32 weeks: large<br>bitemporal horn cysts<br>mild unilateral LVSP | M | 43964 | serology | Livebirth, CMV negative |
| 8 | M | 11 - 15 | serology | Live infant, CMV<br>positive, well | M | 7-16w | serology | Livebirth, CMV positive |
| 9 | F | unknown | Abn US | TOP | F | -2+2w | serology | Livebirth, CMV negative |
| 10 | F | 0-12 | serology | Live infant, CMV<br>positive, clinically<br>well, mild fetal MRI<br>changes | F | 0-12 | serology | Livebirth, CMV negative |
| 11 | M | 10-6 | serology | Live infant, CMV<br>positive, well | M | 0-12 | serology | Livebirth, CMV negative |
| 12 | F | unknown | Abn US | Fetal death | F | 4-6 | serology | Livebirth, CMV negative |
| 13 | F | unknown | serology | Not available | F | NA | Abn US: abnormal<br>Dopplers | NA |

Abn US, abnormal ultrasound; CMV, cytomegalovirus; GA, gestational age; LVSP, lenticulostriate vasculopathy; MRI, magnetic resonance
imaging; NA, not available; SNHL, sensorineural hearing loss; TOP, termination of pregnancy

Of the 13 controls with negative amniotic fluid testing for CMV, three infants had positive urine CMV PCR results at birth, and two infants did not have newborn CMV testing results available. Of the 3 CMV positive infants who had CMV negative amniocentesis results, all were clinically well. One infant had a small subependymal cyst on neonatal brain ultrasound, but was otherwise asymptomatic. Long-term follow up after hospital discharge was not available on live infants.

### Results of RNAseq differential gene expression analysis

At least 30 million reads were obtained per sample. After mapping, quality control and filtering, and quantification of gene expression, the median effective library size was approximately 4 million reads per sample. Due to the random size and relatively short length of cfRNA fragments, most of the read pairs overlapped and there was also substantial adapter contamination. After adapter trimming, some reads were too short for mapping and were excluded. A significant number of reads were also excluded due to likely PCR duplication. There was also evidence that many reads that failed to map were derived from ribosomal RNA.

The initial comparison of the CMV negative group to the CMV infected group using *limma*, with pairing information taken into account, resulted in 210 upregulated and 17 downregulated genes at FDR < 0.05. Ingenuity Pathway Analysis was performed on the 500 most significant DEGs as ranked by FDR. The top canonical pathway enriched in the gene set was interferon signalling (p= 1.58E-10), and the top diseases and disorders were ‘organismal injury and abnormalities, ‘immunological disease, connective tissue disorders and inflammatory disease’.

The Ingenuity Pathway Analysis (IPA) predicted an increase in the disease processes ‘Progressive neurological disorder’ (z-score 2.540, P value < 0.001), as well as inflammatory demyelinating disease (activation score 2.63 p< 0.001). The genes that were identified through the IPA prediction algorithm for progressive neurological disorder are listed in table 4. Necrosis of kidney was also predicted to be increased (activation score 3.161, p< 0.001).

Given the nature of the cfRNA samples and the variability observed, a further analysis was performed using *edgeR*, in combination with *RUVseq*, to estimate and account for additional, unknown sources of unwanted variation present in the data. Compared with the CMV-negative group, this analysis identified 309 up-regulated and 32 down-regulated genes in the CMV-infected group, at FDR < 0.05 (Figure 1). One-hundred and ninety of the statistically significant genes at FDR < 0.05 overlapped between the *RUVseq* and *limma* analyses; and of the top 500 most significant genes ranked by p-value, 373 were the same. Furthermore, the top 78 ranked genes, were the same for both analyses and only differed in their ordering. However, there were 45 statistically significant genes identified by the *RUVseq* analysis that were not in the top 500 ranked genes from the *limma* analysis. The top 5 GO categories associated with these genes were “immune system process”, “immune response”, “response to external biotic stimulus” and “response to biotic stimulus”, suggesting that the *RUVseq* analysis identified additional genes relevant to the biology of CMV infection. Gene list and expression levels of the 12 most statistically significant upregulated genes in the CMV-infected group identified by the *RUVseq* analysis are shown in Figure 2. These genes are involved in interferon signalling and immune defence against viruses. GSEA also showed significant enrichment of multiple GO categories involving the innate immune response to viral infection and interferon signalling (Table 2).

**Figure 1.**
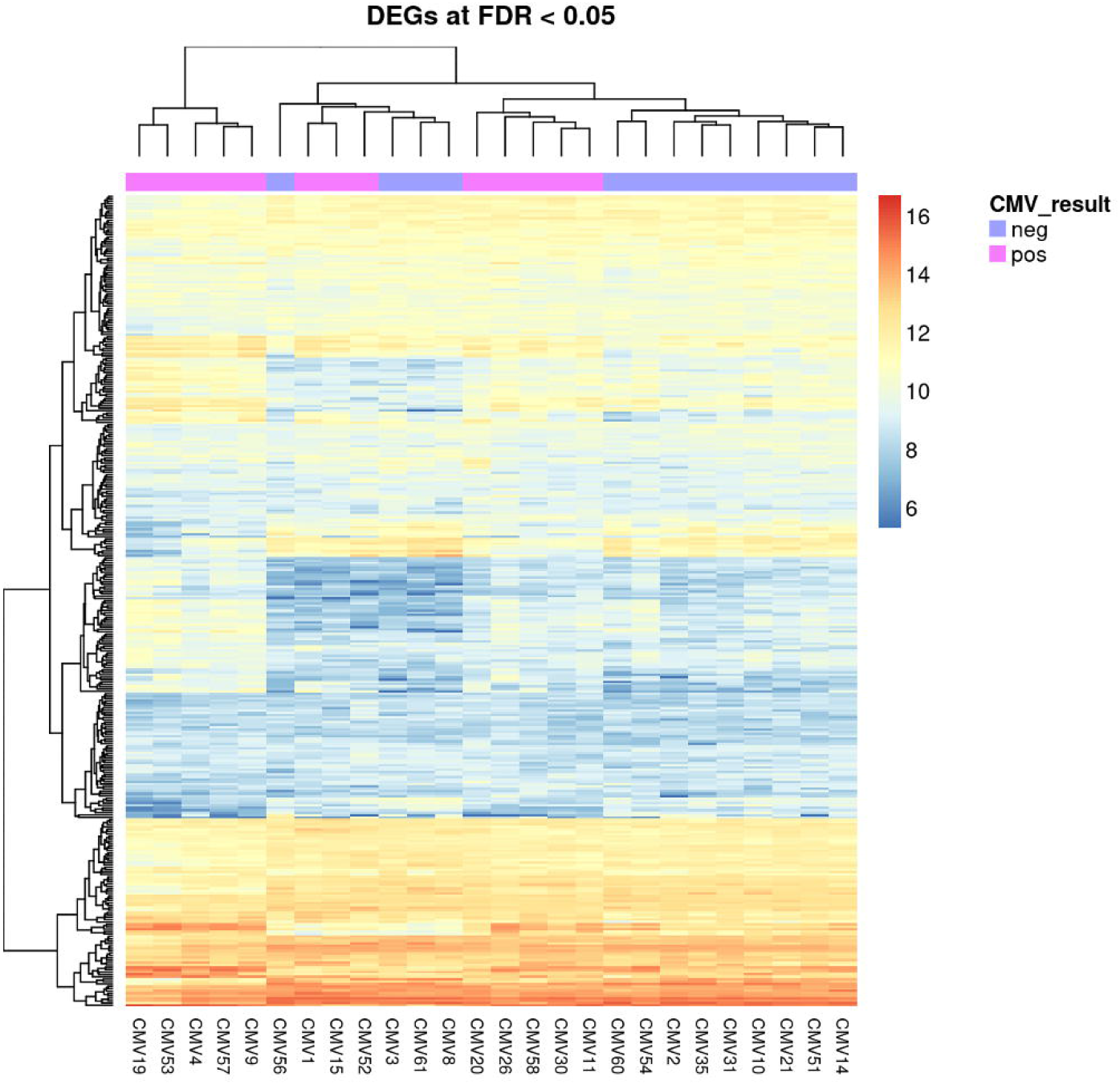
Heatmap of differentially expressed genes in 13 matched cases and controls^1^ ^1^The values depicted are log_2_ counts per million of the *RUVseq* normalised counts. The differentially expressed genes (DEGs) were statistically significant at false discovery rate (FDR) < 0.05.

**Figure 2.**
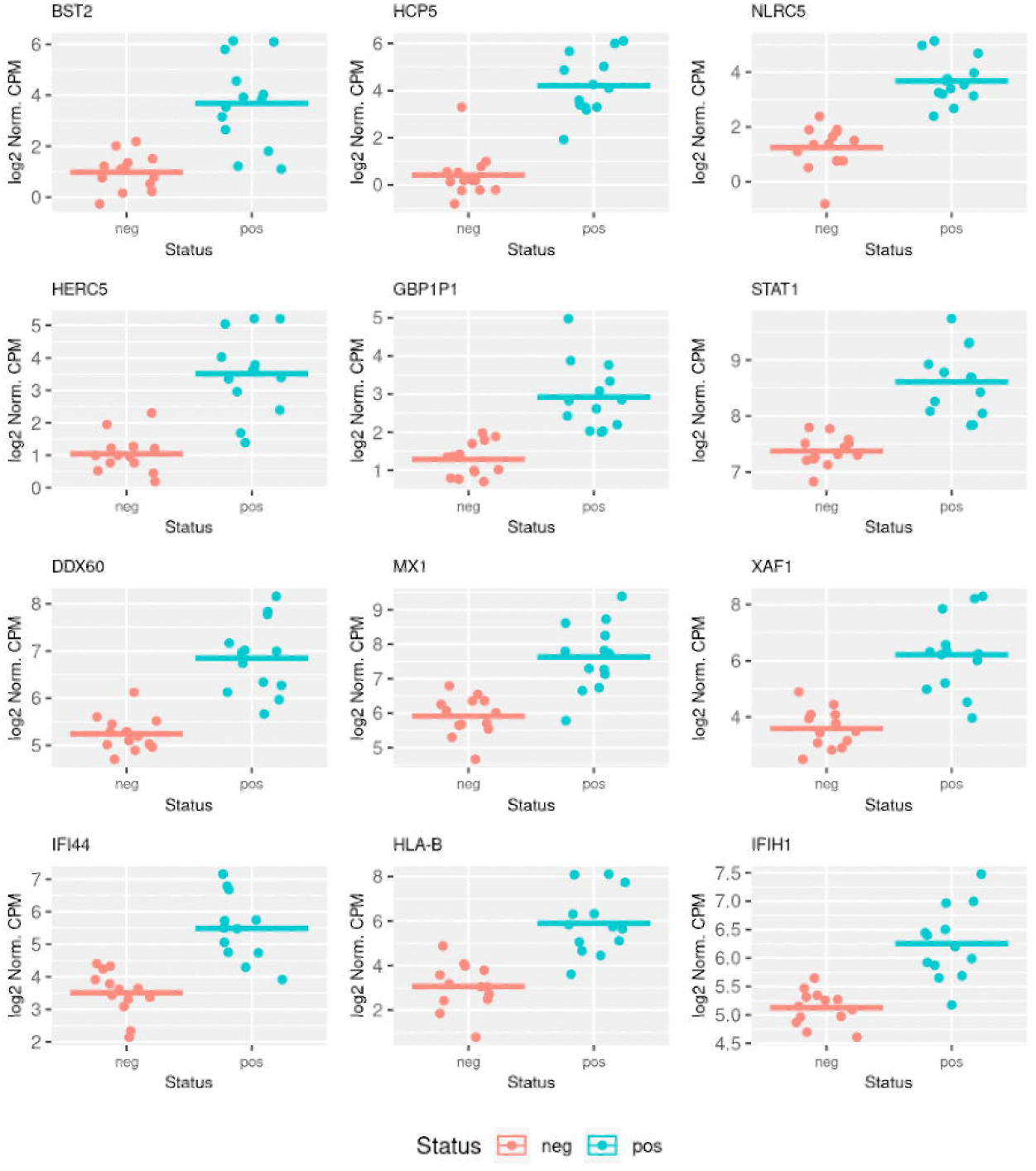
Expression level of the 12 most statistically significant differentially expressed genes in the CMV-positive fetuses^1^ ^1^The values depicted are log_2_ counts per million of the RUVseq normalised counts. BST2, Bone Marrow Stromal Cell Antigen 2; HCP5, HLA Complex P5; NLRC5, NLR Family CARD Domain Containing 5; HERC5, HECT and RLD Domain Containing E3 Ubiquitin Protein; GBP1P1, Guanylate Binding Protein 1 Pseudogene 1; STAT1, Signal Transducer and Activator of Transcription 1; DDX60, DExD/H-Bos Helicase 60; X1, MX Dynamin Like GTPase 1; XAF1, XIAP Associated Factor 1; IFI44, Interferon Induced Protein 44; HLA-B, Major Histocompatibility Complex, Class I, B; IFIH1, Interferon Induced with Helicase C Domain 1.

**Table 2.** Top 20 gene ontology terms significantly enriched in the CMV-infected group

| Gene Ontology Category | Gene Ontology term | No. of genes in gene ontology category | No. of differentially expressed genes in CMV infected fetuses | p value | False discovery rate |
| --- | --- | --- | --- | --- | --- |
| GO:0043207 | response to external biotic stimulus | 849 | 90 | 5.074108e-31 | 1.047702e-26 |
| GO:0051707 | response to other organism | 849 | 90 | 5.074108e-31 | 1.047702e-26 |
| GO:0009607 | response to biotic stimulus | 876 | 91 | 1.048043e-30 | 2.163789e-26 |
| GO:0045087 | innate immune response | 516 | 70 | 1.932626e-30 | 3.989906e-26 |
| GO:0006955 | immune response | 1177 | 105 | 4.709784e-30 | 9.722877e-26 |
| GO:0098542 | defense response to other organism | 628 | 74 | 3.480584e-28 | 7.184970e-24 |
| GO:0006952 | defense response | 913 | 88 | 3.201128e-27 | 6.607769e-23 |
| GO:0034340 | response to type I interferon | 74 | 29 | 9.011344e-27 | 1.860032e-22 |
| GO:0071357 | cellular response to type I interferon | 70 | 28 | 3.497598e-26 | 7.219042e-22 |
| GO:0060337 | type I interferon signalling pathway | 70 | 28 | 3.497598e-26 | 7.219042e-22 |
| GO:0002376 | immune system process | 1887 | 127 | 4.838293e-25 | 9.985269e-21 |
| GO:0051607 | defense response to virus | 178 | 38 | 8.177496e-24 | 1.687590e-19 |
| GO:0009615 | response to virus | 231 | 41 | 2.183999e-22 | 4.506901e-18 |
| GO:0044419 | interspecies interaction between<br>organisms | 1439 | 102 | 3.558582e-21 | 7.343134e-17 |
| GO:0009605 | response to external stimulus | 1674 | 111 | 6.533592e-21 | 1.348141e-16 |
| GO:0002252 | immune effector process | 776 | 70 | 8.031013e-20 | 1.657039e-15 |
| GO:0034097 | response to cytokine | 802 | 70 | 4.991354e-19 | 1.029816e-14 |
| GO:0019221 | cytokine-mediated signalling pathway | 505 | 53 | 7.088885e-18 | 1.462508e-13 |
| GO:0034341 | response to interferon-gamma | 121 | 26 | 1.082392e-16 | 2.232974e-12 |
| GO:0071345 | cellular response to cytokine stimulus | 740 | 63 | 1.372889e-16 | 2.832132e-12 |

CMV, cytomegalovirus

Of the 32 significantly down-regulated genes in the CMV-infected group, eight were neurodevelopmental genes with known roles in brain development and preferentially expressed by brain (Table 3). Six of 16 host restriction factors known to target human CMV had significantly upregulated gene transcripts in the CMV-infected group (*LGAL9, PML, SP100, MX2, BCL2L14, RSAD2*).

**Table 3.**
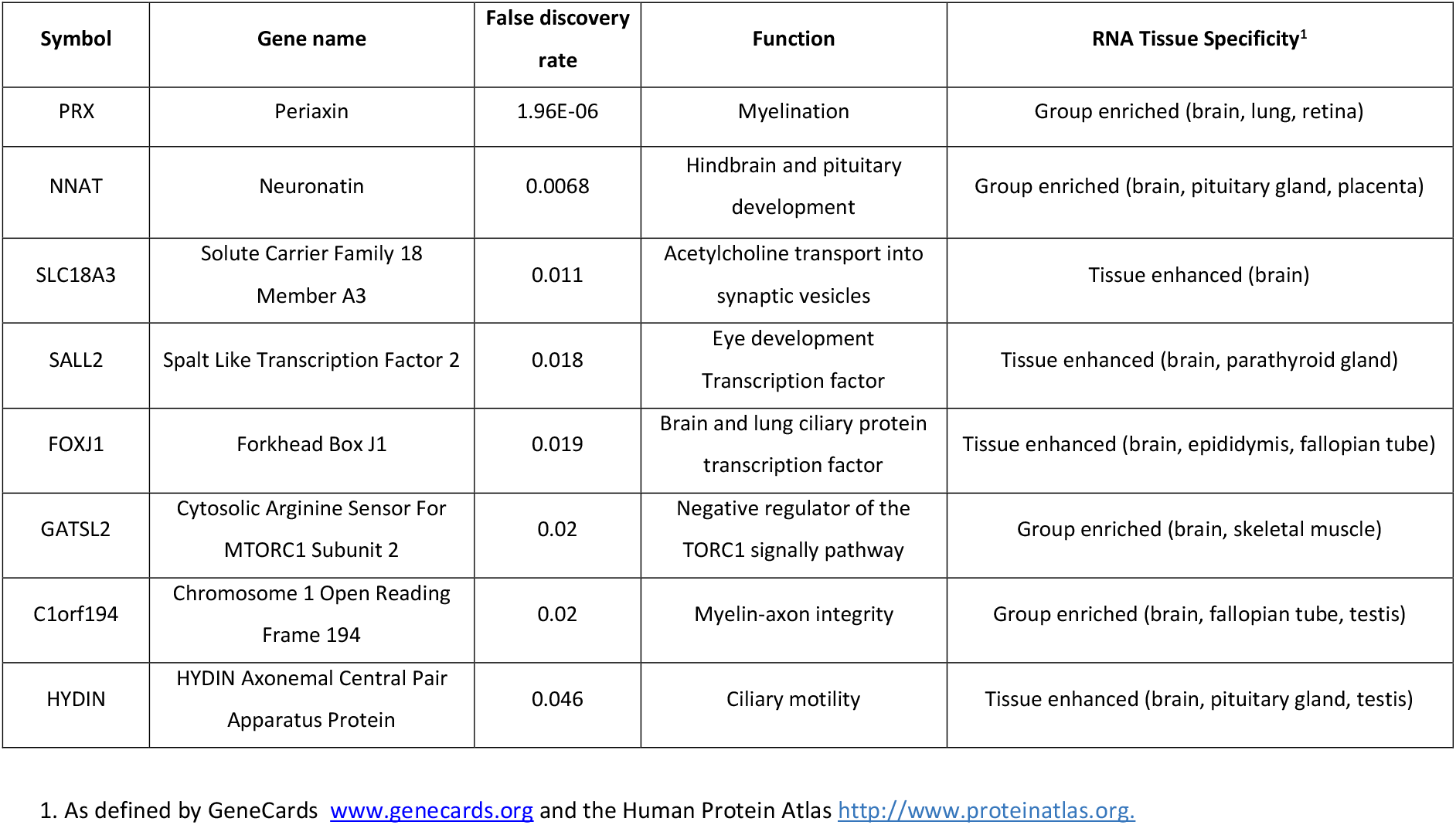
Neurodevelopmental genes significantly downregulated in the CMV-infected group

**Table 4.** Ten genes identified by Ingenuity Pathway Analysis to predict progressive neurological disorder among CMV-infected fetuses.

| Gene symbol | Gene name | Predicted | Function UniProtKB/Swiss-Prot Summary |
| --- | --- | --- | --- |
| RSAD2 | Radical S-Adenosyl Methionine Domain Containing 2 | Increased | Interferon-inducible iron-sulfur (4FE-4S) cluster-binding antiviral protein which plays a major role in the cell antiviral state induced by type I and type II interferon. |
| OAS3 | 2'-5'-Oligoadenylate Synthetase 3 | Increased | Interferon-induced, dsRNA-activated antiviral enzyme which plays a critical role in cellular innate antiviral response. In addition, it may also play a role in other cellular processes such as apoptosis, cell growth, differentiation and gene regulation. |
| MX1 | MX Dynamin Like GTPase 1 | Increased | Interferon-induced dynamin-like GTPase with antiviral activity against a wide range of RNA viruses and some DNA viruses. |
| CASP1 | Caspase 1 | Increased | Signaling pathways of apoptosis, necrosis and inflammation |
| IRF7 | Interferon Regulatory Factor 7 | Increased | Key transcriptional regulator of type I interferon (IFN)-dependent immune responses and plays a critical role in the innate immune response against DNA and RNA viruses. |
| ISG15 | ISG15 Ubiquitin Like Modifier | Increased | Ubiquitin-like protein which plays a key role in the innate immune response to viral infection |
| PSEN1 | Presenilin 1 | Increased | Catalytic subunit of the gamma-secretase complex, an endoprotease complex that catalyzes the intramembrane cleavage of integral membrane proteins such as Notch receptors and amyloid-beta precursor protein. Involved in the regulation of neurite outgrowth |
| OAS1 | 2'-5'-Oligoadenylate Synthetase 1 | Increased | Interferon-induced, dsRNA-activated antiviral enzyme which plays a critical role in cellular innate antiviral response. In addition, it may also play a role in other cellular processes such as apoptosis, cell growth, differentiation and gene regulation. |
| IFIT1 | Interferon Induced Protein With Tetratricopeptide Repeats 1 | increased | Interferon-induced antiviral RNA-binding protein acting as a sensor of viral single-stranded RNAs and inhibiting expression of viral messenger RNAs |
| DDC | Dopa Decarboxylase | decreased | Involved in neurotransmitter metabolism. Catalyzes the decarboxylation of L-3,4-dihydroxyphenylalanine (DOPA) to dopamine, L-5-hydroxytryptophan to serotonin and L-tryptophan to tryptamine. |

### Analysis results after excluding CMV-infected fetuses with ultrasound abnormalities

We repeated the *RUVseq* differential gene expression analysis in 10 pairs after excluding the three CMV cases with ultrasound abnormalities at recruitment. With this smaller sample size, there were fewer statistically significant DEGs (n=63), but with a similar overall expression profile compared with the total cohort. The top 10 genes remained unchanged as did the significant GO categories that involved interferon signalling and innate immune response to viral infection. Of the significantly down-regulated neurodevelopmental genes in table 3, only *periactin* had FDR < 0.05 after excluding cases with US abnormalities.

### Differential exon usage

Comparison of differential exon usage between infected and non-infected samples revealed 17 differentially used exons arising from 13 genes with FDR < 0.1. When exons belonging to multiple genes were excluded, five genes with differential exon usage remained (Supp. Table 1). After repeat analysis following the exclusion of the three CMV cases with ultrasound abnormalities at recruitment, there were eight genes identified with differential exon usage, including one gene, *ALDOB* with two exons altered between infected and non-infected samples (Supplemental file 2). Some of these genes were well recognised as having alternative splicing.

## Comment

### Principal findings

In this next generation sequencing study, we revealed novel features of the human intraamniotic immune response and fetal neurodevelopment associated with CMV infection. Functional interpretation of the DEGs supported our hypothesis that inflammation and neurological disease would be the most dysregulated biological processes in the CMV-infected group.

The most upregulated genes in our cCMV cohort were involved in the innate immune response to viral infection in both the GSEA and the IPA functional analyses. At the level of individual genes, we noted increased expression of multiple cellular restriction factors known to target CMV. These restriction factors play various roles in the host immune response against CMV: Galectin-9 restricts entry of CMV into host cells, *MXB* interferes with the nuclear delivery of CMV genomes, *PML* nuclear bodies associate with CMV to induce epigenetic silencing, and *BCL2L14* restricts CMV viral replication via regulation of the type I interferon response.^13^ *RSAD2* codes for the cytoplasmic antiviral protein viperin that is directly induced by CMV.^14^ Our results thus provide the first human data on the involvement of these specific restriction factors in congenital CMV infection. We assume that the gene expression related to the innate immune response reflect a predominately fetal response, since our control group included women with evidence of maternal, but not fetal, CMV infection during pregnancy. Neurohistochemistry studies have reported that second trimester human fetuses exhibit an innate immune response driven by the microglia.^3^ However, we recognise that immune cells of both maternal and fetal origin are detectable in amniotic fluid,^15,16^ so it is possible that both maternal and fetal immune systems contribute to the intraamniotic response observed in cCMV.

Our results also provide insights into the pathogenesis of brain injury in human fetuses. CMV brain pathogenesis has been most extensively studied in experimental settings, such as *in vitro* human CMV infection of neural progenitor cells or *in vivo* murine CMV infection.^17^ It has been proposed that CMV disrupts neuronal migration and synapse formation through several mechanisms, including direct cytopathic effects of viral replication.^18^ In our study, the most downregulated genes in the CMV-infected group included genes related to neuronal cell differentiation (*NNAT*), myelination (*PRX*), and synaptic function (*SLC*), providing the first human data to complement the results of experimental mouse and neural stem cells models.^19,20^

### Implications for fetal medicine

We specifically targeted recruitment at the time of clinically-indicated amniocentesis in fetuses without ultrasound abnormalities as proof of principle that differential gene expression can be detected prior to the manifestation of structural brain anomalies. Through this process, potential mRNA biomarkers could be selected. However, the clinical validation of biomarkers that predict outcome after congenital CMV would require much larger cohorts with a range of fetal and postnatal phenotypes, and long-term neurodevelopmental outcomes. In this era, where routine serological screening in pregnancy is being proposed^21^, the inevitable rise in diagnostic testing that would company such a change in practice may facilitate a sufficient sample size to validate this approach. A predictor of neurological outcome – perhaps in combination with imaging and other diagnostic features – may help future counselling of parents about postnatal outcomes. Our data provides candidate biomarkers that await further studies and potential external validation by independent researchers.

### Implications for translational research

The AF transcriptome was first comprehensively described in 2012.^8^ It has since been explored by several independent groups, confirming its potential for tracking fetal and placental function throughout gestation.^10–12^ The majority of published studies of the AF transcriptome have been performed with gene expression microarrays. More recently, RNAseq has been employed to tackle some of the technological challenges of working with low abundance samples of cfRNA.^12,15,22^ However, the putative sources of the gene transcripts are only directly inferred based on known patterns of tissue-specific gene expression.

The strength of our study is the use of next generation sequencing to better understand the pathophysiology of CMV infection in live second trimester fetuses, which has previously been limited by the paucity of suitable animal or *in vitro* models. We optimized the biological validity of our findings by using a paired design, controlling for sex and gestation, which are known to influence fetal gene expression. Unlike other RNAseq studies, we avoided using pooled samples as this has been shown to identify DEGs with high false positivity rates and low positive predictive values in RNAseq experiments.^23^

Our results were likely impacted by the relatively low effective read library size, due to technical challenges inherent to the sequencing of AF cfRNA. Based on our experience, future AF RNAseq studies should endeavour to minimise adapter contamination by assessing the sizing profile of the final libraries and selecting the sequencing read length based on the shortest insert length, and ensuring that the selected read length is shorter than the average insert size. Although PCR duplication is known to introduce potential biases and duplicate removal is warranted, incorporation of unique molecular identifiers (UMIs) could help to optimise duplicate removal in RNAseq of AF cfRNA.^24^ Ensuring efficiency of ribosomal RNA removal is also paramount and selection of the most appropriate method should be guided by the aims of the study.

Although we reached our prespecified minimum target sample size, many of the DEGs did not meet our stringent criteria for statistical significance after excluding the three fetuses with ultrasound abnormalities. As the yield of amniotic fluid cfRNA is known to be insufficient for performing both RT-qPCR and RNAseq on the same samples, we intentionally used a matched pair design with sufficient biological replicates to ensure confidence in our results. Although we did not have sufficient RNA to perform technical validation with qRT-PCR, multiple studies have shown a high consistency between RNA-seq and qRT-PCR results.^25^ Ideally, we would have validated the DEGs in an independent cohort. However, during the course of this study, clinical practice has evolved to include the offer of fetal therapy with valaciclovir^26^ to prevent fetal infection following maternal primary infection in first trimester, which was an exclusion criterion for our study. We therefore decided not to extend the study period as we anticipated very low recruitment rates. However, future studies including participants using valaciclovir therapy may assist in assessing the fetal response to antiviral therapy.

## Conclusion

Here, in this next generation sequencing study, we reveal new insights into the pathophysiology of congenital CMV infection in live human fetuses. These data on the upregulation of the intraamniotic innate immune response to CMV infection and the downregulation of neurodevelopmental genes may inform future approaches to developing prognostic markers and assessing fetal responses to in utero therapy.

## Data Availability

All analysis code presented in this manuscript can be found at https://jovmaksimovic.github.io/amnio-cell-free-RNA/index.html. The analysis website was created using the workflowr (1.6.2) R package.27 The GitHub repository associated with the analysis website is at: https://github.com/JovMaksimovic/amnio-cell-free-RNA.
Ethical approval precluded deposition of the sequencing datasets in a public genomic data repository; however the de-identified, processed, unnormalized read counts may be available to researchers upon reasonable request to, subject to Mercy Health ethics committee approval.

https://github.com/JovMaksimovic/amnio-cell-free-RNA

## Acknowledgements and funding

This study was funded by a Sylvia Charles Viertel Clinical Investigator grant and the National Health and Medical Research Council (#1196010 and #1105603 to LH and #1146128 to NJH). The funding sources had no involvement in the study design, collection, analysis, data interpretation, or writing of the report.

We thank the laboratory staff at the Centre for Human Genetics, UZ Leuven, and the biobank staff at the Department of Obstetrics and Gynecology, University of North Carolina, for assistance in sample collection and transport.

## Declarations

The authors have no conflicts of interest to declare.

## Data sharing

All analysis code presented in this manuscript can be found at https://jovmaksimovic.github.io/amnio-cell-free-RNA/index.html. The analysis website was created using the *workflowr* (1.6.2) R package.^27^ The GitHub repository associated with the analysis website is at: https://github.com/JovMaksimovic/amnio-cell-free-RNA.

Ethical approval precluded deposition of the sequencing datasets in a public genomic data repository; however the de-identified, processed, unnormalized read counts may be available to researchers upon reasonable request to, subject to Mercy Health ethics committee approval.

## Supplemental files

Supplemental File 1. Detailed statistical methods.

Supplementary File 2. Table S1. Genes exhibiting statistically significant (FDR < 0.1) differential exon usage between CMV-infected cases and controls

## Notes

**Disclosure statement** The authors report no conflicts of interest.

### Competing Interest Statement

The authors have declared no competing interest.

### Author Declarations

Prospective ethical approval for this study was obtained from human research ethics committee of Mercy Health (R16-24) on 27 June 2016.

