## Supplemental file 1 for "RNAseq of amniotic fluid cell-free RNA: insights into the pathophysiology of congenital cytomegalovirus infection"

**Supplemental data 1. Detailed statistical methods.**

*Sample size calculation*

Our sample size calculation took into account estimates of: depth of sequencing, coefficient of variation, magnitude of differential expression detected, false positive rate and power. Assuming an average read depth of 20 million reads/sample, equal within group coefficient of variation of 0.4, an effect size of 2 fold, 0.05 false positive rate and 80% power, the number of subjects required per group was seven.^1^ This target was in keeping with recommendations for RNA-seq experiments^2^ and other published studies using amniotic fluid.^3,4^ As the yield of RNA from amniotic fluid is known to be low, we aimed to exceed the minimum sample size to optimise the number of biological replicates. We therefore aimed for a target sample size of 12 matched pairs, with a minimum of eight matched pairs.

*Differential gene expression analysis*

Paired-end RNA sequencing was performed on the Novaseq 6000 at 30 million reads per sample. Reads were assessed for quality using FastQC (v0.11.5) and adapters and poor-quality bases trimmed using Trimommatic (v0.39)^5^; single reads with pairs deemed too short for mapping were retained. Paired and single-end reads were mapped to the human genome (hg38) separately using STAR (v2.7.5b)^6^ and potential PCR duplicates, as indicated by Picard (v2.17.3), were discarded. Read counts were summarised across genes using featureCounts from Subread (v1.5.2).^7^ GENCODE (v34) gene annotations were used for junction-mapping and read counting. Data quality and processing metrics were summarised using MultiQC (v1.8).^8^ Paired-end and single-end counts for the same sample were summed for downstream analysis.

Two approaches were used to identify differentially expressed genes (DEGs). Initially, the “voomWithQualityWeights” function was used to estimate the mean-variance relationship of log_2_-counts per million transformed data.^9^ This generated observational-level weights as well as sample-specific quality weights, which were incorporated into the linear models fitted for each gene using *limma*.^10^ DEGs between CMV positive and CMV negative samples were identified using empirical Bayes moderated t-tests, taking pairing information into account.^11^ To protect against hypervariable genes, robust empirical Bayes shrinkage of the gene-wise variances was used.^12^ The Benjamini-Hochberg method was employed to adjust for multiple testing.^13^ Genes with a false discovery rate (FDR) < 0.05 were considered statistically significant. Ingenuity Pathway Analysis (IPA) using IPA Version 9.0 software (Ingenuity, Redwood City, CA) was then applied to the results.

Alternatively, to account for the high variability inherent in cfRNA data, the “RUVg” function from the *RUVseq* package^14^ was used to perform factor analysis to estimate the unwanted variation present in the data with human house-keeping genes designated as negative controls.^15^ Following dispersion estimation, generalized log-linear models were fitted to the counts for each gene using *edgeR*^16^, taking into account pairing information and the first component of unwanted variation estimated by *RUVseq*. DEGs were identified using likelihood ratio tests. As for the *limma* analysis, the Benjamini-Hochberg method was used to adjust for multiple testing and genes with an FDR < 0.05 were considered statistically significant.

All functional analyses, other than IPA, were performed on the results of the *RUVseq* analysis. Testing for DEGs was performed with and without CMV cases with fetal ultrasound abnormalities and their matched controls.

*Functional analysis*

DEGs were interpreted using the “goana” function from the *limma* R/Bioconductor package. “Goana” was used to test for over-representation of GO terms amongst the statistically significant DEGs, adjusting for gene length bias.^17^ Multiple testing adjustment was performed using the Benjamini-Hochberg method. GO terms with an FDR < 0.05 were considered statistically significant. .

The RNA tissue specificity of the downregulated genes were assessed using the Human Protein Atlas to identify neurodevelopmental genes.^18^ We defined a gene as ‘neurodevelopmental’ if it was shown to have enhanced expression in the central nervous system (CNS) in the Human Protein Atlas and had a known neurological function as annotated in the public resource GeneCards.^19^

Functional analyses were performed using IPA Version 9.0 software (Ingenuity, Redwood City, CA). Ingenuity identifies over-represented biological processes in a given dataset and calculates a significance score for each result using the right tailed Fisher’s test. Ingenuity Pathway Analysis was performed on the 500 most significant DEGs from the *limma* analysis as ranked by FDR. IPA downstream effects analysis was used to predict the activation or inhibition of specific processes on the basis of the direction of differential regulation of genes. Results were considered statistically significant if z-score >2 (activation in CMV-infected fetuses) or less than 2 (inhibition in CMV-infected fetuses). Results pertaining to cancer were excluded as these are known to be confounded in fetal datasets due to normal cell proliferation during development.^20,21^ Sixteen cellular restriction factors known to be involved in the human immune response to CMV infection were also individually examined for evidence of significant differential expression.^22^

*Differential exon usage analysis*

Alternative splicing allows a single gene to code for multiple proteins by producing transcripts comprised of different exons; this process may be dysregulated in disease. Differential exon usage (DEU) is present when there are differences in the relative proportion of exons in the transcripts between two groups. Reads were processed, mapped and duplicates removed as previously described. The GENCODE (v34) gene annotation was prepared for exon counting with HTSeq^23^ using the “dexseq_prepare_annotation.py” script provided with the *DEXSeq* R/Bioconductor package. Paired and single-end reads were then counted separately using HTseq (v0.11.2) executed via the “dexseq_count.py” script. Paired-end and single-end counts for the same sample were summed for downstream analysis. We performed exploratory analysis for DEU between the CMV-infected and the control group by fitting generalised linear models to each known exon using the *DEXSeq* package.^24^ Exons with a Benjamini-Hochberg adjusted p-value < 0.1 were considered statistically significant.

**Data sharing**

All analysis code presented in this manuscript can be found at https://jovmaksimovic.github.io/amnio-cell-free-RNA/index.html. The analysis website was created using the *workflowr* (1.6.2) R package.^25^ The GitHub repository associated with the analysis website is at: https://github.com/JovMaksimovic/amnio-cell-free-RNA.

Ethical approval precluded deposition of the sequencing datasets in a public genomic data repository, however the de-identified, processed, unnormalized read counts may be available to researchers upon reasonable request to.
