## Supplemental File 2 for "RNAseq of amniotic fluid cell-free RNA: insights into the pathophysiology of congenital cytomegalovirus infection"

| **Symbol** | **Gene name** | **Function** | **Exon ID^1^** | **Log_2_FC** | **Adjusted p-value** | **US abn included** |
| --- | --- | --- | --- | --- | --- | --- |
| MDFIC | MyoD Family Inhibitor Domain Containing | Transcriptional regulation of viral genome expression | E017 | 0.1186993 | 0.0016686 | No |
| SERPINA1 | Serpin Family A Member 1 | Serine protease inhibitor | E016 | -0.3616374 | 0.0323076 | Yes |
| CTPS1 | CTP Synthase 1 | Enzyme that converts UTP to CTP | E030 | -1.6987830 | 0.0386619 | Yes |
| C1orf116 | Chromosome 1 Open Reading frame 116 | Putative androgen-specific receptor | E001 | 0.1454078 | 0.0456537 | No |
| SERPINA1 | Serpin Family A Member 1 | Serine protease inhibitor | E016 | -0.3426304 | 0.0480108 | No |
| ALDOB | Aldolase, Fructose-Bisphosphate B | Glycolytic enzyme | E020 | -0.5227151 | 0.0616827 | No |
| ALDOB | Aldolase, Fructose-Bisphosphate B | Glycolytic enzyme | E023 | -0.7271615 | 0.0616827 | No |
| IFT122 | Intraflagellar Transport 122 | Cilia formation during neuronal patterning | E012 | -1.3888995 | 0.0662329 | Yes |
| ALDOB | Aldolase, Fructose-Bisphosphate B | Glycolytic enzyme | E020 | -0.5309091 | 0.0662329 | Yes |
| ZG16 | Zymogen Granule Protein 16 | Putative role in protein trafficking | E004 | -0.4264955 | 0.0716148 | No |
| DENR | Density Regulated Re-Initiation and Release Factor | Translation initiation | E015 | 0.2620617 | 0.0967841 | No |
| KIAA0100 | KIAA0100 | Putative role in membrane trafficking | E041 | -0.5139657 | 0.0967841 | No |
| FAM111A | FAM111 Trypsin Like Peptidase A | Maintenance of genomic integrity | E027 | 0.2656931 | 0.0967841 | No |
| PITRM1 | Pitrilysin Metallopeptidase 1 | Degradation of mitochondrial transit peptides | E024 | -1.0108556 | 0.0986710 | Yes |

Supplemental file 2. Genes exhibiting statistically significant (FDR < 0.1) differential exon usage between CMV-infected cases and controls
